# Clinical multi-omics reveals the role of Tuomin Zhiti Decoction Intervention in Allergic Rhinitis from the perspective of biological network

**DOI:** 10.1101/2024.03.10.24303911

**Authors:** Weibo Zhao, Boyang Wang, Lingyao Kong, Qi Wang, Shao Li

## Abstract

**Background:** Increasing evidence showed that seasonal allergic rhinitis (SAR), as an allergy disease, could be alleviated with traditional Chinese medicine (TCM) formula, one example being Tuomin Zhiti Decoction (TZD). However, as a complex composition of TCM herbs, the mechanism of TZD in the treatment of SAR remain unclear.

**Purpose:** Uncover the mechanism of TZD for treating SAR based on computational analysis and clinical multi-omics experiments.

**Study design:** Integrate computational analysis including network target analysis and machine learning algorithms with clinical multi-omics experiments and public omics data.

**Methods:** By analyzing TZD’s composition through a network-based method, we identified the biological effects of each compound, constructing a comprehensive biological network to elucidate TZD’s molecular and pathway mechanisms against AR. Single-arm clinical trials on the gut microbiome and serum transcriptomics corroborated our computational insights. Further validation through public omics data highlighted key TZD compounds, paving the way for future research.

**Results:** TZD was discovered to exert a regulatory effect on various modules associated with AR, as demonstrated by the constructed biological network. Insights from gut microbiome and serum transcriptomics in clinical trials, where immune-related microbiomes represented by Prevotella, as well as pathways and biological processes including antigen processing and presentation, activation and regulating immune cell surface receptor, were markedly enriched (*P* value < 0.05), indicated that TZD played a pivotal role in modulating immune processes and the immune cells against AR. With the verification of multi-omics, it was determined that TZD potentially influences immune responses and downstream immune cells like CD4+ T cells, through both direct mechanisms involving antigen and indirect mechanisms mediated by gut microbiomes in the treatment of AR.

**Conclusion:** Through combination of computational prediction and analysis together with clinical multi-omics, a network target based framework provided a new insight for uncovering the mechanism of TZD for treating AR.

## Introduction

Allergic Rhinitis (AR) is an inflammatory disease of the nasal mucosa, mediated by allergen-specific Immunoglobulin E (IgE). Its clinical manifestations include sneezing, nasal itching, runny nose, and nasal congestion^1^. Severe cases can lead to olfactory dysfunction and various complications. Epidemiological studies have revealed a global incidence rate of approximately 40% for allergic rhinitis, with a particularly high prevalence rate of 19.1% in North China among Chinese adults^2^. These findings indicate that allergic rhinitis is not only a widespread issue but also a growing medical concern. While it may not be life-threatening, the recurrent symptoms and associated complications significantly impact patients’ quality of life and mental well-being. Given its high incidence and detrimental effects on patients, allergic rhinitis is increasingly recognized as a global health problem^1,2^.

The common symptom-controlling treatments include nasal corticosteroids, antihistamine, and allergen immunotherapy^3^. These treatments, while recommended by the official guidelines, may not be an optimal solution for AR patients in the long term^1^.In recent years, more and more patients are looking for complementary and alternative medicines to treat AR, such as Chinese herbal medicine^4^. A growing number of studies have produced promising results on the curative effect of traditional Chinese medicine (TCM) treatment on AR, all with relatively few side effects^5–8^, such as TZD. Tuomin Zhiti Decoction (TZD) is a formula made up of eleven different herbs, including Magnoliae Flos (Xinyi), Xanthii Fructus (Cangerzi), Mume Fructus (Wumei), Cicadae Periostracum (Chantui), Saposhnikoviae Radix (Fangfeng), Ganoderma (Lingzhi), Astragali Radix (Huangqi), Scutellariae Radix (Huangqin), Lilii Bulbus (Baihe), Glycyrrhizae Radix et Rhizoma (Gancao) and Nidus Vespae (Fengfang). One animal studie have demonstrated that TZD could decrease the AR-induced damage to the nasal mucosa, and clearly ameliorate nasal symptoms through reducing OVA-specific immunoglobulin E (IgE) and histamine release^9^. However, TZD’s effect has not been clarified yet.

As a combination of network science, artificial intelligence and multi-omics, network target was proposed by Li^10–12^ to uncover the complex role of TCM in the treatment of diseases. From the perspective of systemic biology and medicine^13,14^, network target has been continuously developing and succeeded in many aspects of TCM researches, including revealing mechanism of TCM^15^, discovering biomarkers^16^ and determining efficacy compounds^17,18^.

Therefore, this study evaluates the clinical efficacy and safety of TZD in patients with AR, aims to uncover the mechanism of TZD against AR and provides high-quality evidence to support its clinical application on the basis of network target analysis and clinical multi-omics.

## Methods and Materials

### Prediction of biological effect profiles of TZD

In order to estimate the molecular level function of TZD and its compositions, DrugCIPHER algorithm^19^ was performed to make prediction on genome-wide biological effect profiles for each compound in TZD. In addition, the compounds recorded in each herb of TZD was obtained from HERB database^20^. Then, a statistical strategy proposed in previous study^21^ was used to calculate the holistic targets of every herb in TZD, as well as TZD itself. Holistic targets with adjust *P* value less than 0.05 (Benjamini-Hochberg adjustment) were remained for further analysis.

### Enrichment analysis of AR and TZD

Molecules related to AR were collected in two ways, including curated ones from CTD database^22^ and ones predicted by a text-mining algorithm Gendoo ^23^. Furthermore, Kyoto Encyclopedia of Genes and Genomes (KEGG) and Gene Ontology (GO) enrichment analysis was performed based on the biomolecules related to AR as well as holistic targets of herbs in TZD and biological effect profiles of compounds in TZD. KEGG pathways and GO terms with adjust *P* value less than 0.05 were kept for further use. In addition, KEGG pathways were classified into different categories based on the class and sub class pre-defined.

### Modular network construction of TZD against AR

In order to depict the modular regulatory role of TZD against AR, modular biological network was constructed based on the molecular level and pathway level prediction results. Significantly enriched pathways and biological processes were classed into five modules depending on their roles in AR, including Allergic Reaction and Inflammatory Response, T Cell Immunity and Differentiation, Immune Cell Mobility and Response, Phagocytosis and Antigen Presentation, as well as Cell Signaling and Communication. Interactions between herbs and targets, pathways or biological processes, were based on the predicted holistic targets and the enrichment result, respectively.

### Clinical study and Trials Design

This study conducted a 12-week, single-arm clinical trial involving 80 patients with allergic rhinitis (AR) who received TuominZhiti (TZD) intervention. The study adhered to the principles of the Declaration of Helsinki, obtained approval from the Ethics Committee of Shanghai Dongfang Hospital (No.), and was registered in the Chinese Clinical Trial Registry (ChiCTR) under registration number ChiCTR-IOR-1. Prior to enrollment, all participants provided informed consent. Clinicians recorded all clinical data, while efficacy and safety were evaluated through relevant clinical phenotypes measured and analyzed by the Clinical Laboratory of BUCM at baseline and after 12 weeks of intervention. Blood samples from each participant were collected following standard procedures and stored at -80℃ within two hours for subsequent multi-omics detection.

### Detection of intestinal flora structure and abundance by 16SrRNA gene sequencing

Bacterial DNA extraction from fecal samples: fecal samples were suspended in sterile 0.1moL/L sodium phosphate buffer (PBS), vortexed for 30 min, and centrifuged at 200g for 3 times, 5min each time, to remove coarse particles and collect the supernatant; centrifuged at 9000g for 3min, collected the precipitate, washed, resuspended, and stored at -70°C for subsequent DNA extraction. Bronchial lavage fluid was centrifuged, precipitated, resuspended, and stored for DNA extraction. Total DNA was extracted by Bead beating method. All DNA samples were stored at -20°C for subsequent PCR analysis.

## Results

### Biological effect profiles of TZD

To uncover the mechanism of TZD in the treatment of AR and build the macro-micro correlation models to construct relationship between phenotypes and genotypes, based on our previous algorithm DrugCIPHER^19^, the biological effect profiles of each compound belonging to herbs in TZD that recorded in HERB database^20^ were predicted and processed. With the validation of PubMed, the biological effect profiles of representative compounds in TZD were found to be highly covered by literature reports (Figure 1A). Furthermore, we predicted the holistic targets of herbs in TZD according to a statistical model proposed in our previous study^21^, as well as the holistic targets of TZD. It could be found that different herbs in TZD shared some common compounds as well as common targets (Figure 1B, C). These chemical composition and computational prediction results to some extent demonstrate the potential synergistic effects among various herbs in the intervention of AR by TZD, which aligns with the TCM theory “Jun-Chen-Zuo-Shi” (JCZS).

**Figure 1.**
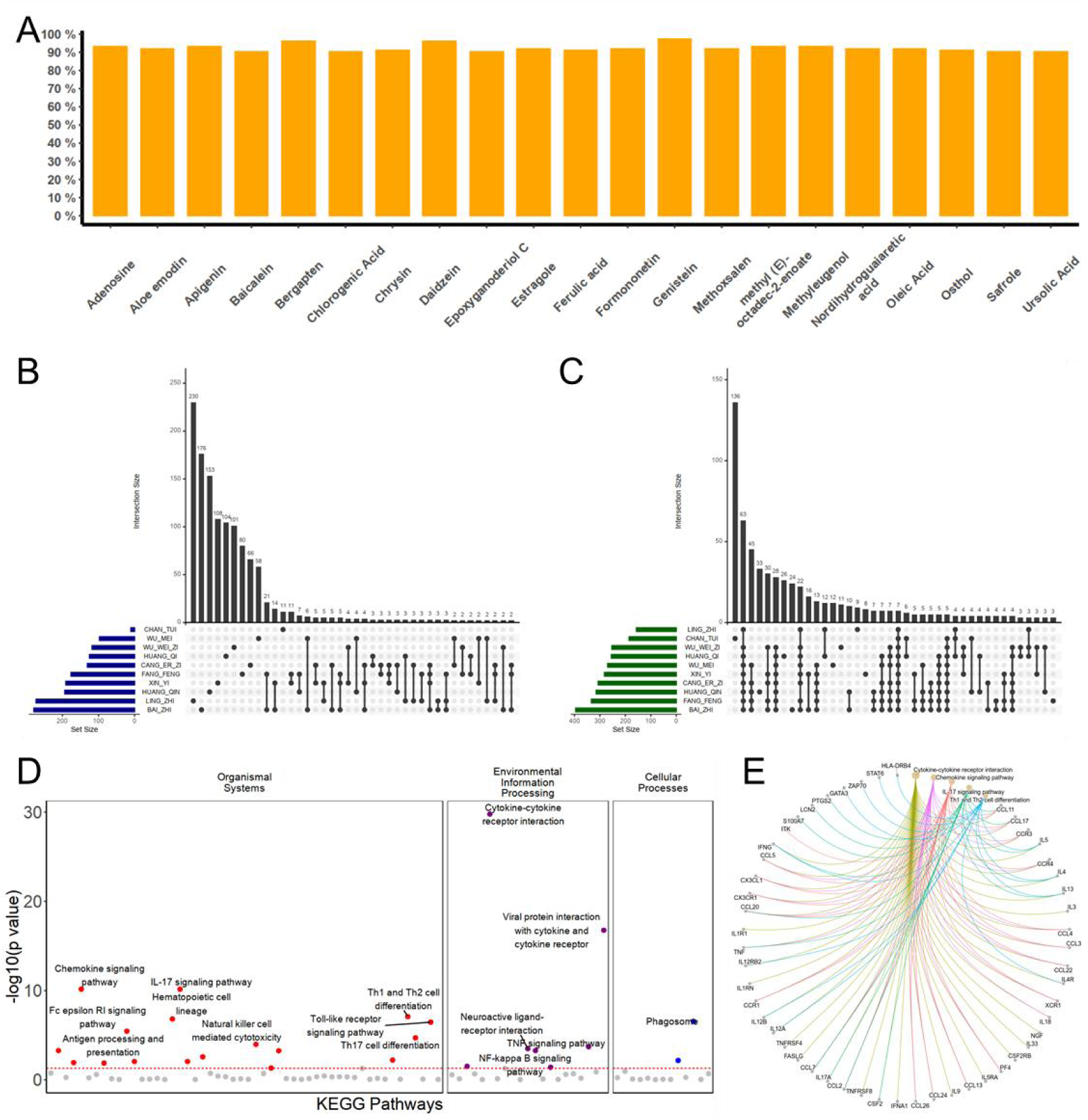
(A) Bar plot showing the literature cover rate of predicted biological effect profiles of some representative compounds in herbs of TZD. (B) and (C) Upset graphs showing shared compounds or predicted targets of different herbs of TZD, respectively. (D) Dot plot showing enriched pathways of collected molecules related to AR and their classification. (E) Network of top ranked pathways enriched by collected molecules related to AR, corresponding molecules in these pathways and their relationships.

### Network target analysis of TZD against AR

In order to depict the potential intervention effects of TZD in treating AR, we collected AR-related molecules from CTD database^22^ as well as a disease-gene predicting method Gendoo^23^, and conducted analyses based on these molecules. Pathways related to these molecules were primarily concentrated in Organismal Systems module dominated by the immune system and Environmental Information Processing module represented by Signaling Transduction. In addition, some pathways related to cellular processes were also significantly enriched (Figure 1D, E). Certain molecules in these pathways were reported to play important role in AR. For example, IL-4, IFN-γ, and TNF-α were found to increase mucosal permeability and blocking IL-4 helped maintain the integrity of the mucosal barrier and prevented the reduction of tight junctions^24^. Cytokines such as IL-1 and TNF-αare commonly present in the body’s inflamed areas and these mediators play a key role in inflammation by recruiting additional inflammatory cells, triggering the release of more inflammatory agents, and activating afferent nerves, thereby amplifying the inflammatory response^25^. Type 2 cytokines (IL-4, IL-5, and IL-13) along with innate cytokines (IL-25, IL-33, and TSLP) were also recognized as significant biomarkers, indicating their crucial role in various allergic biological processes and allergic diseases^26^.

With the combination of pathology of AR and potential effect of TZD, it was found that the holistic targets of TZD fell in five classification including Metabolism, Genetic Information Processing, Environmental Information Processing, Cellular Processing and Organismal Systems (Figure 2A). Among them, the regulation of TZD on sub classifications Signal Transduction and Immune System might play the most critical role in the treatment of AR, as the holistic targets of TZD located mostly on pathways related to these sub classifications. Focusing on the pathway level, we discovered that TZD exerts its therapeutic effect on AR through a modular regulation mechanism, including Allergic Reaction and Inflammatory Response, T Cell Immunity and Differentiation, Immune Cell Mobility and Response, Phagocytosis and Antigen Presentation, as well as Cell Signaling and Communication (Figure 2B). In these modules, pathways like Th1 and Th2 cell differentiation were reported to play dominant roles in AR^27,28^, which also served as important intervention targets in the treatment of AR. A shift Th1/Th2 balance towards IL-4 (Th2) has been linked to the onset of allergic and atopic diseases, like AR^29^. In addition, expression of Fc epsilon RI is recognized not only in the context of atopic dermatitis but is broadly associated with various allergic diseases, indicating its key role in the immune response related to allergies^30^. These pathways were also of vital importance in the targets network (Figure 2C, D). Besides, biological processes associated with IFN-γ were reported closely to allergic diseases represented by AR^31–33^, as well as those related to cell adhesion^34–36^ and T cell^37–39^, which aligned with their significant positions within the multi-layered biological networks (Figure 2E, F).

**Figure 2.**
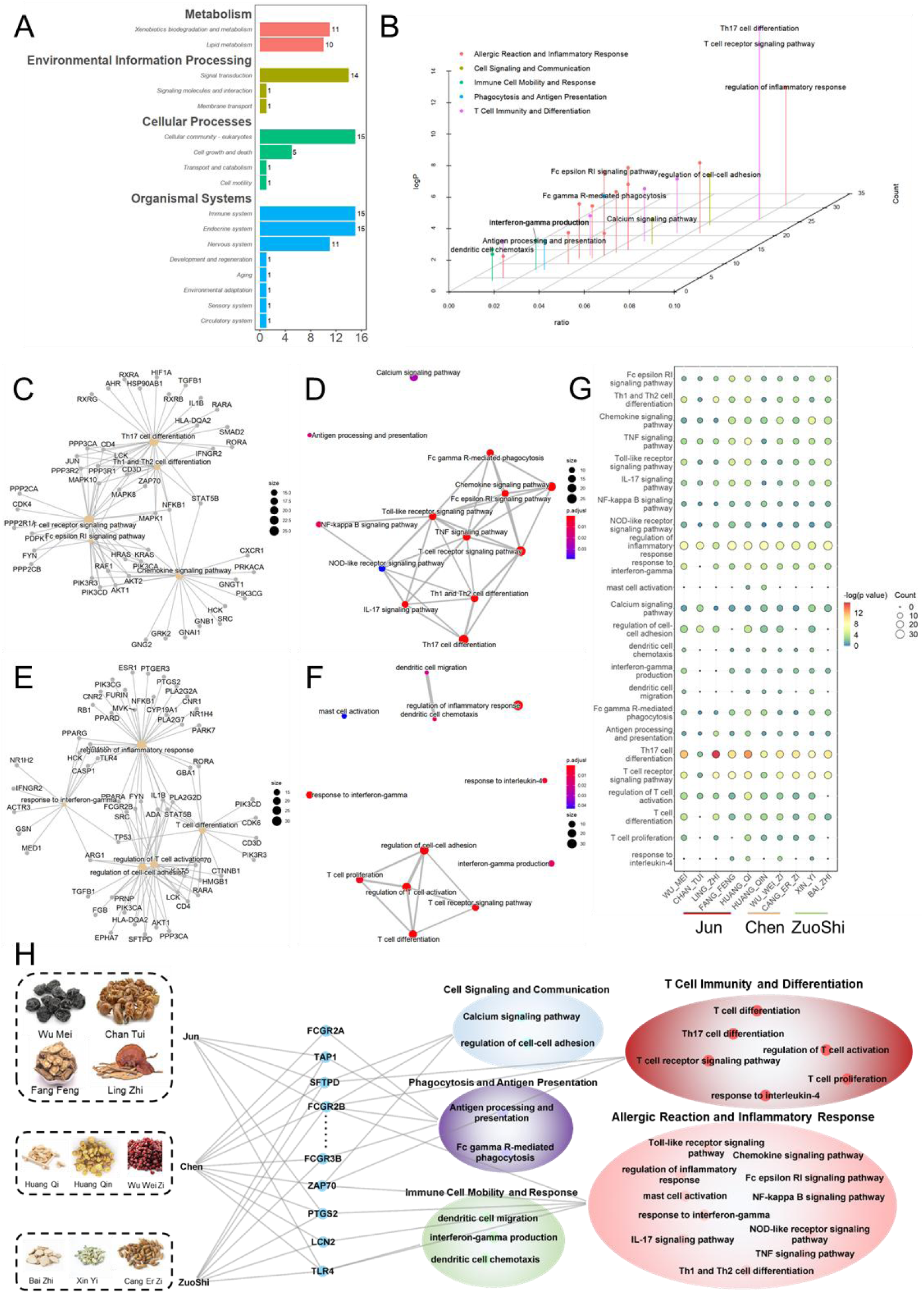
(A) Bar plot of the distributions of holistic targets of TZD in the treatment of AR categorized in different classifications and sub classifications. (B) 3D dot plot of pathways or biological processes representing modular mechanism of TZD against AR and their assigned modules. (C) Molecule-pathway relationship network showing associations between certain holistic targets of TZD and pathways in modular mechanism of TZD against AR. (D) Pathway-pathway network showing the associations between pathways in modular mechanism of TZD against AR. (E) Molecule-biological processes relationship network showing associations between certain holistic targets of TZD and biological processes in modular mechanism of TZD against AR. (F) Biological processes-biological processes network showing the associations between biological processes in modular mechanism of TZD against AR. (G) Dot plot showing the enrichment result of pathways or biological processes in modular mechanism of TZD against AR for herbs regarded as Jun, Chen or Zuoshi in TZD. (H) Multi-layer network composed of herbs, molecules, modules and pathways or biological processes showing the network regulatory effect of TZD in the treatment of AR.

### Potential roles of different herbs in TZD

As it is widely acknowledged that the composition of Chinese formula follows certain compatibility principles, it is crucial to predict the role of each herb in TZD in the intervention of AR. The enrichment levels of different Chinese herbs in TZD on pathways and biological processes related to the modular mechanism of TZD in intervening AR (Figure 2G). Among these pathways or biological processes and their related pharmacological effects, there were certain reports associating them with herbs in TZD. For example, Jun herb Wu Mei, could significantly decrease the expression level of IL-17^40^ and formula composed of it were reported to have influence on Th1 cell related cytokines like IFN-γ^41^ and Th2 cell related cytokines like IL-4^42^. In addition, Ling Zhi had been shown to boost the Th1 immune response, resulting in elevated levels of IFN-γ and IL-2^43,44^ and reduce levels of IL-1β, IL-6, and TNF-α^45^. And Fang Feng could regulate the release of NO, TNF-α, IL-1β, and IL-6, as well as the density of immune cells like macrophages^46^.

Finally, we constructed a multi-layer biological network representing the effect of TZD against AR (Figure 2H). This network was built on the basis of enrichment results for every herb of TZD in the modular pathways or biological processes. And Jun herbs were predicted to take effect mainly on Allergic Reaction and Inflammatory Response module together with T Cell Immunity and Differentiation module, while Chen herbs potentially affects all the five modules and might exerted a relatively comprehensive regulatory effect. And herbs regarded as Zuoshi, intended to regulate Phagocytosis and Antigen Presentation module and Immune Cell Mobility and Response module according to prediction and analysis. On the molecular level, JCZS herbs played synergistic regulatory roles on these AR-related molecules which acted important jobs in these modules.

### Clinical Trials demonstrating the effectiveness of TZD against AR

A 12-week single-arm clinical trial was conducted to evaluate the efficacy of TZD intervention in patients with AR (Figure 3A). In this study, TZD significantly relieved both nasal and eye symptoms of SAR, including running noses, nasal itching, nasal congestion, ocular redness/itching etc. The results confirmed the significant clinical benefits of TZD in alleviating symptoms, including a notable reduction in TNSS, TESS, and IgE levels, as well as a substantial improvement in RQLQ (quality of life) for AR patients (*P* value <0.05; Figure 3B, C). Long-term follow-up revealed that during the second year and in 2022 (one year after intervention), the total score for AR was 5.88, which showed statistical significance compared to pre-intervention scores (*P* value < 0.05). In addition, TZD treatment was found to be safe given that most patients reported no adverse reactions.

**Table1.**
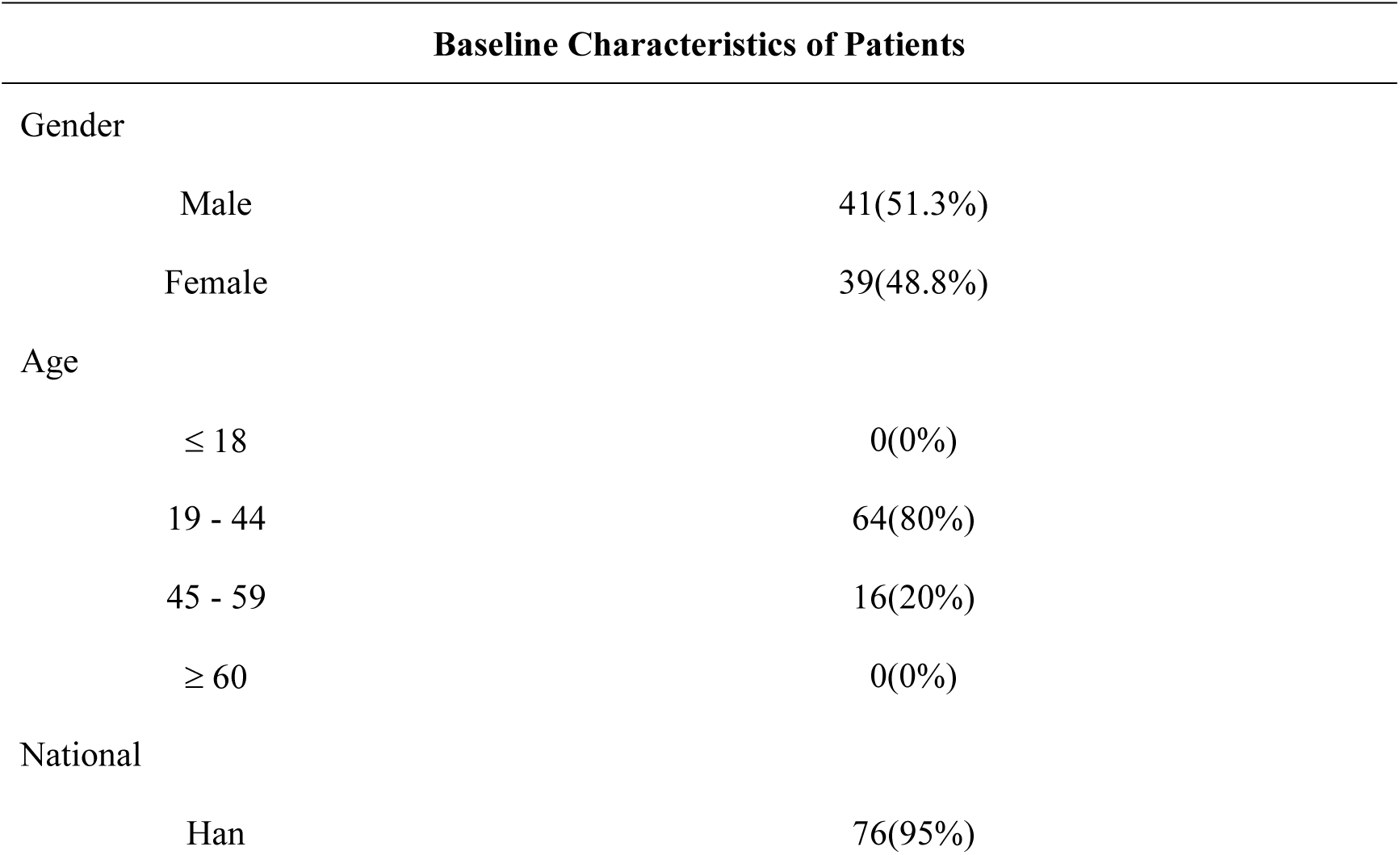
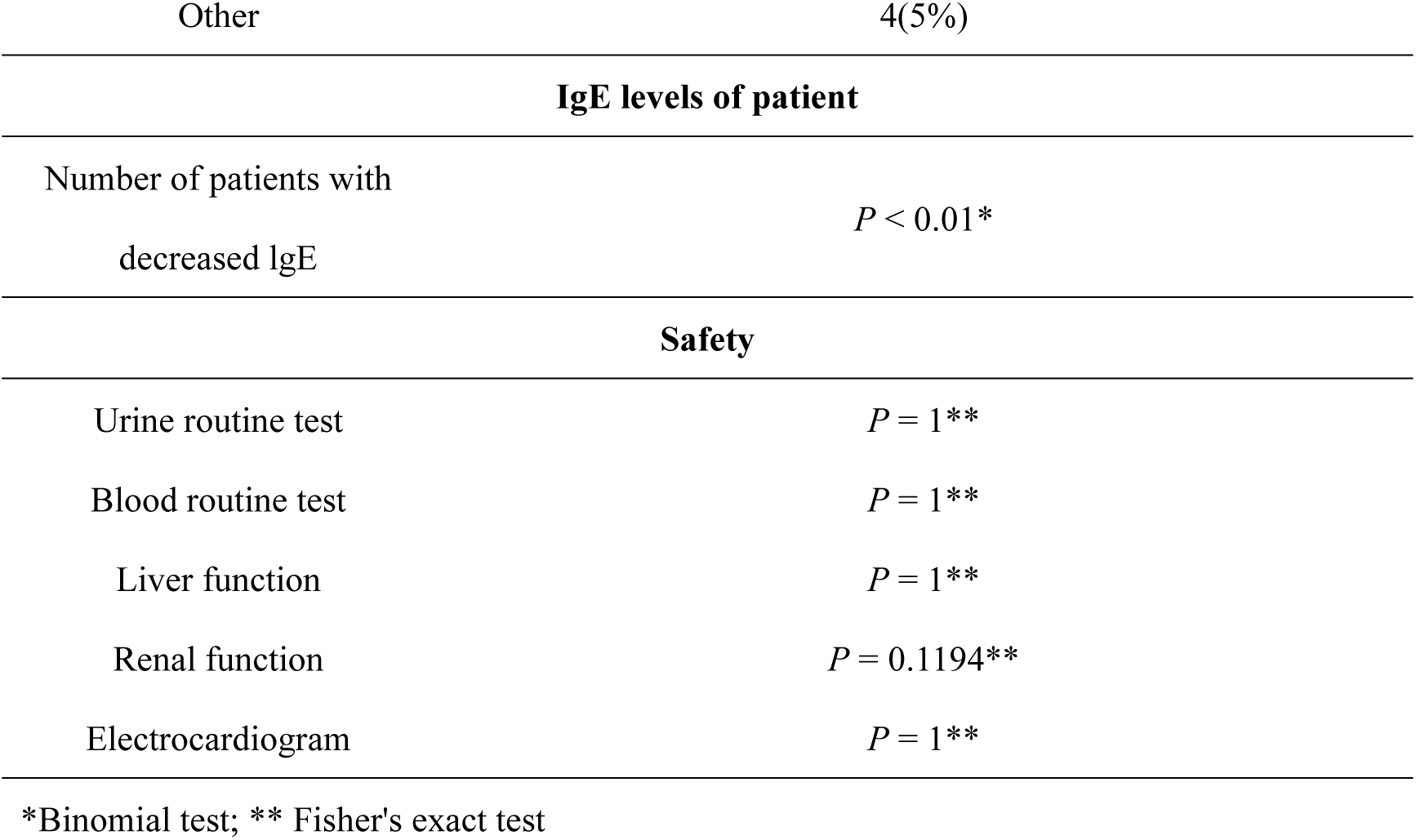
Clinical trials of TZD against AR.

| Baseline Characteristics of Patients |  |  |
| --- | --- | --- |
| Gender |  |  |
|  | Male | 41(51.3%) |
|  | Female | 39(48.8%) |
| Age |  |  |
|  | ≤ 18 | 0(0%) |
|  | 19 - 44 | 64(80%) |
|  | 45 - 59 | 16(20%) |
|  | ≥ 60 | 0(0%) |
| National |  |  |
|  | Han | 76(95%) |

|  |  |
| --- | --- |
| Other | 4(5%) |
| **IgE levels of patient** |  |
| Number of patients with decreased IgE | $P < 0.01^*$ |
| **Safety** |  |
| Urine routine test | $P = 1^{**}$ |
| Blood routine test | $P = 1^{**}$ |
| Liver function | $P = 1^{**}$ |
| Renal function | $P = 0.1194^{**}$ |
| Electrocardiogram | $P = 1^{**}$ |

\*Binomial test; \*\* Fisher's exact test

**Figure 3.**
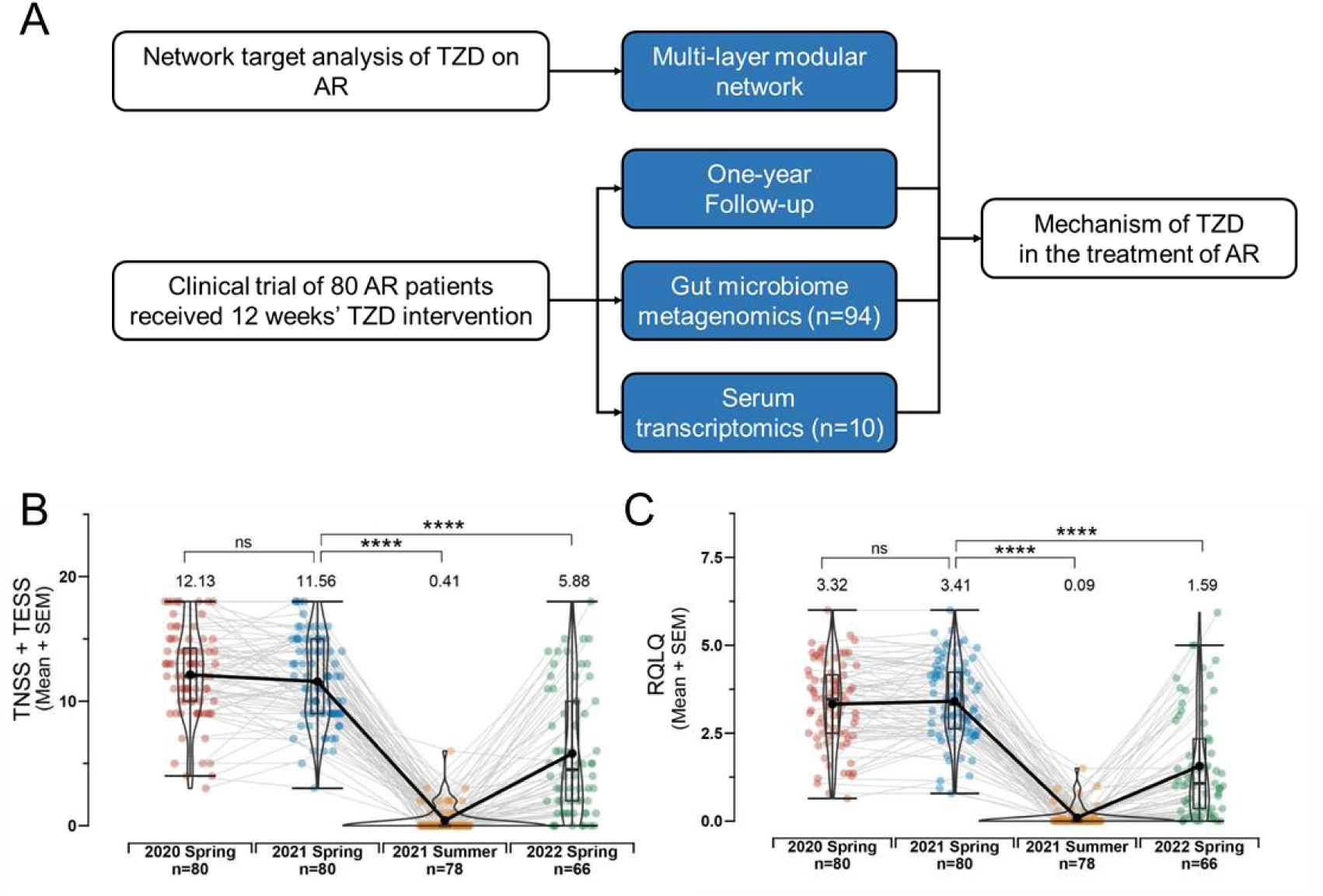
(A) Workflow of overall study process with the combination of computational prediction and clinical trials including multi-omics and follow-up. (B) Clinical statistical results of TNSS + TESS before and after TZD intervention and during follow-up. (C) Clinical statistical results of RQLQ before and after TZD intervention and during follow-up.

To gain insights into the mechanism of action underlying TZD treatment for AR, we further investigated changes in gut microbiota and serum transcriptomics before and after TZD intervention. In total, gut microbiome metagenomics were conducted on 94 samples and serum transcriptomics were conducted on 10 samples.

### Regulatory effect of TZD intervention on gut microbiome

The analysis of gut microbiome metagenomics revealed changes in species relative abundance across various taxonomic levels like phylum and genus. For instance, at the phylum level, the proportion of Firmicutes decreased, while Proteobacteria increased (Figure 4A). And at the genus level, an increase in the proportion of Megamonas and Bacteroides was observed, whereas the proportion of Prevotella_9 and Faecalibacterium decreased after treatment (Figure 4B, C). These shifts indicate significant microbial composition changes due to the treatment. More specifically, detailed statistical analyses were conducted on species with higher abundance in the categories of family, genus, and species (Figure 4D). Significantly changes species like Prevotella and Faecalibacterium were reported to be associated with AR or allergic inflammation^47–49^, as well as key modules in network target of TZD against AR.

**Figure 4.**
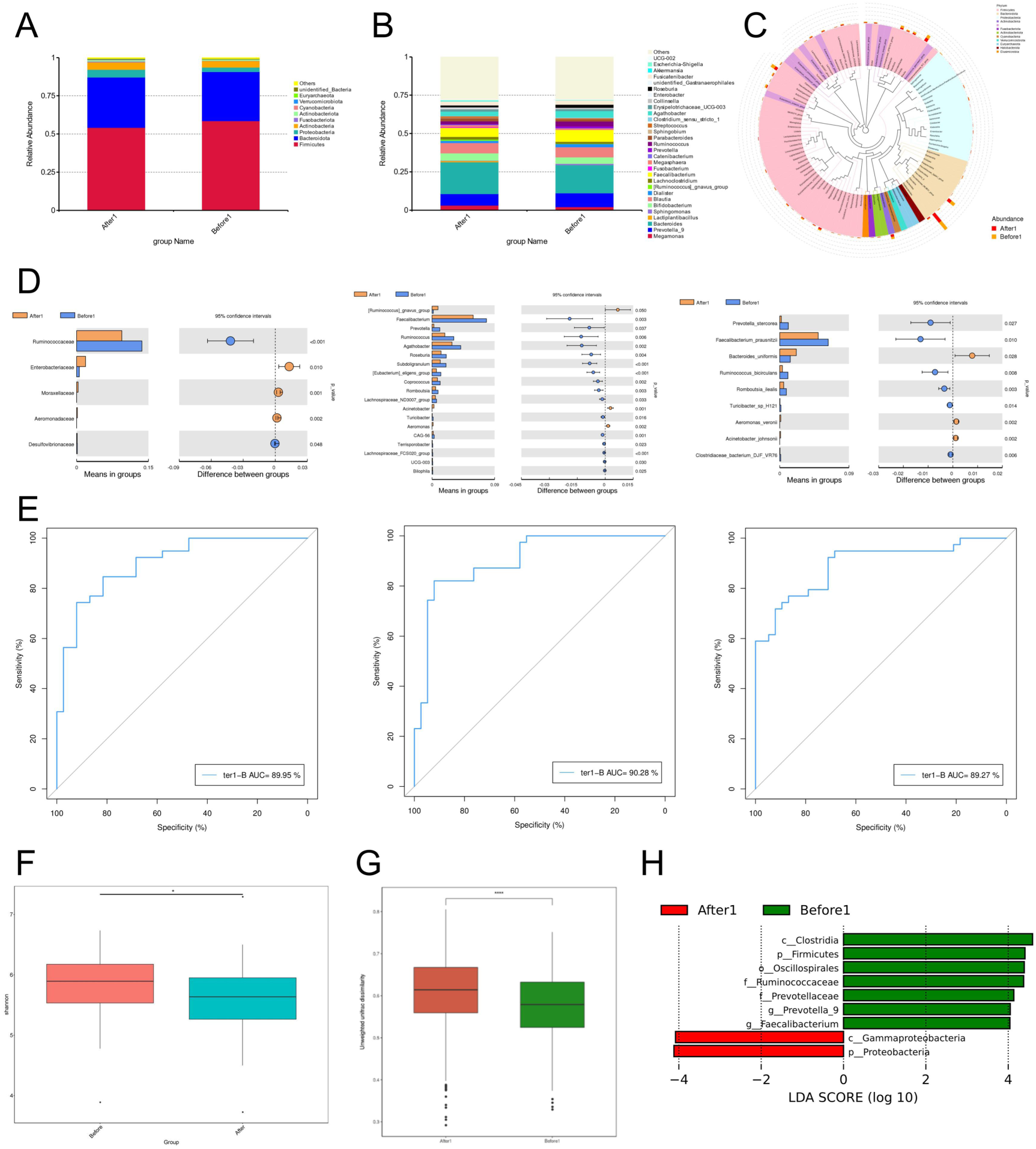
(A) Bar plot showing the proportion of gut microbiome classified in different phyla before and after the intervention of TZD. (B) Bar plot showing the proportion of gut microbiome classified in different genera before and after the intervention of TZD. (C) Classification tree plot of gut microbiome classified in different genera. (D) Significantly changed gut microbiome classified in different familiae, genera and species from left to right, respectively. (E) Random forest AUC score for gut microbiome classified in different familiae, genera and species from left to right, respectively. (F) Box plot of Shannon index showing Alpha diversity before and after the intervention of TZD. (G) Box plot of unweighted unifrac distance showing Beta diversity before and after the intervention of TZD. (H) Bar plot showing LDA score of significantly changed gut microbiome classified in different genera calculated by LEfSe algorithm.

In addition, random forests model trained on the gut microbiomes also showed that species in the categories of family, genus, and species could be appropriate biomarkers for evaluating the efficacy of TZD in the treatment of AR (Figure 4E). Significant differences in the Shannon index of Alpha diversity before and after treatment (*P* value < 0.05) indicate notable changes in microbial abundance and evenness after TZD intervention (Figure 4F). Besides, the Beta diversity, using unweighted UniFrac distances (Figure 4G), revealed significant disparities between the groups before and after TZD intervention (*P* value < 0.001), with a greater variation observed within samples after intervention. Based on LEfSe (LDA Effect Size) analysis, it was found that LDA scores of species like Prevotella were higher in samples before TZD intervention and those of species including Proteobacteria and gamma Proteobacteria were higher in samples after TZD intervention (Figure 4H), which to some extent was consistent with previous statistical results.

### Transcriptomics revealing molecular level changes in serum

We collected serum samples from 5 clinical patients before and after the intervention of TZD and conducted transcriptome sequencing. To focus on the transcriptional level changes in serum molecules before and after the intervention of TZD, R package DESeq2 was performed to obtain differential expressed genes (DEGs). Totally, 26 DEGs were filtered (adjust *P* value < 0.05, log2Foldchange ≥ 1 or ≤ -1) with paired statistical test and kept for further analysis (Figure 5A). Further, based on these DEGs, pathway like antigen process and presentation was significantly enriched (Figure 5B), so as some immune response and natural killer (NK) cell related biological processes. With previous findings, these enriched pathways and biological processes to some extent, suggested that the role TZD played in immune process like antigen process and presentation and those related to immune cells could be an important mechanism in the treatment of AR. According to previous studies, T cells acted as an important role in allergic inflammation in AR^50–52^. Besides, NK cells were also of vital importance in the pathology of AR^53–55^, like resolving eosinophilia and inflammatory crosstalk to eosinophil. Then, Gene Set Variation Analysis (GSVA)^56^ was performed to estimate pathway level changes (Figure 5C) and it was found that apart from antigen process and presentation, many T cell related biological processes were significantly enrich, including T cell cytokine production, activation, proliferation and chemotaxis.

**Figure 5.**
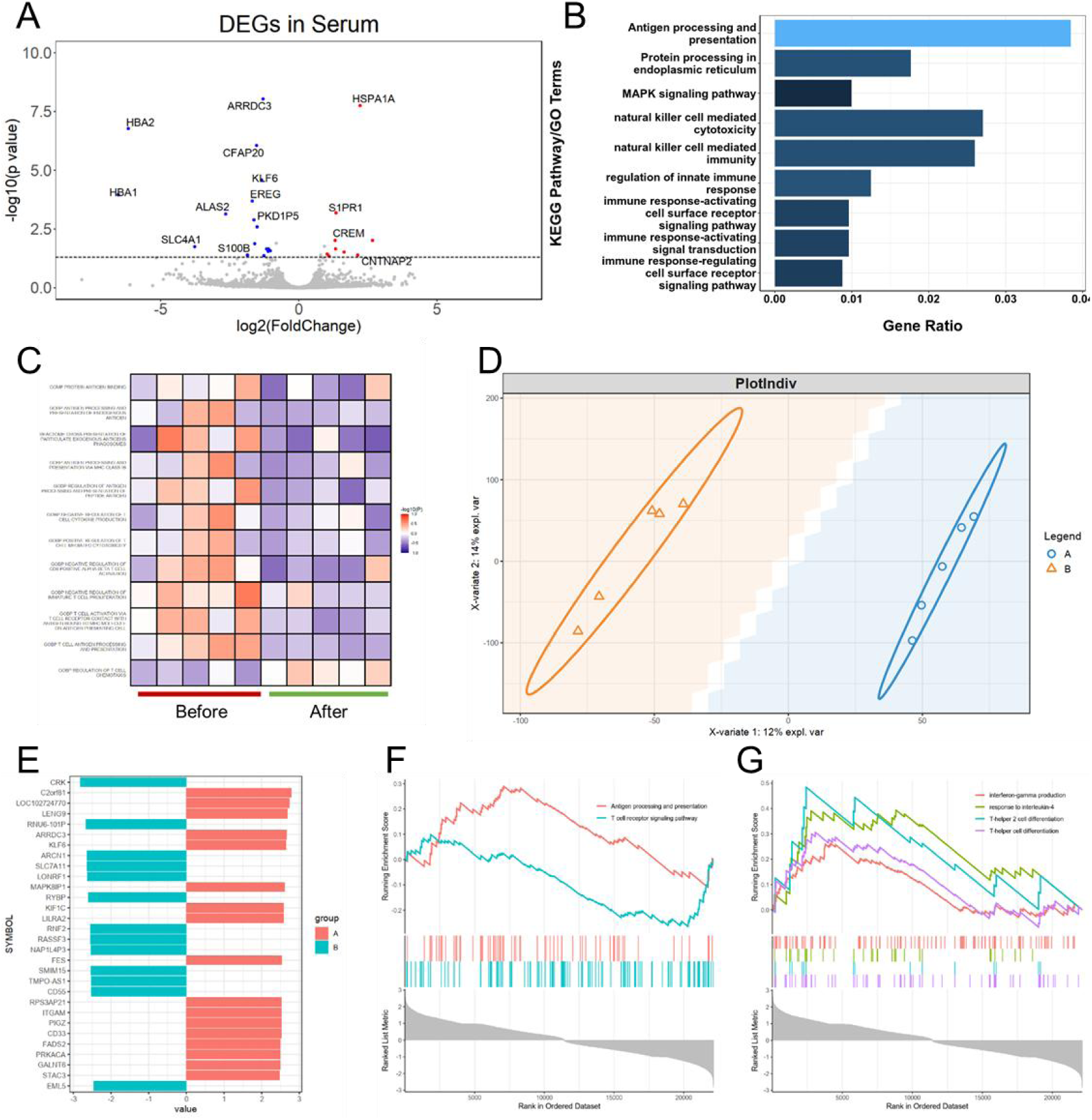
(A) Volcano plot showing DEGs obtained through statistical calculations by DESeq2, with log2 Foldchange as x axis and -log10 adjust p value as y axis. (B) Bar plot of KEGG and GO enrichment results of DEGs. (C) Heatmap showing GSVA score of different gene sets in samples before and after the intervention of TZD. (D) PLS-DA model demonstrating the separability of transcriptomics in a low-dimensional principal component space before and after the intervention of TZD. (E) Bar plot of the directional VIP score of representative genes calculated from PLS-DA model. (F) GSEA result showing the significantly enriched pathways related to antigen process and T cell (*P* value < 0.05). (G) GSEA result showing the significantly enriched biological process related to previous predicted network target of TZD against AR (*P* value < 0.05).

Further, from another perspective, a typical machine learning model, Partial Least Squares Discriminant Analysis (PLS-DA) to make deeper analysis. Based on the contributions of various features to the first two principal components, it was easy to see that samples before and after intervention are divided into two categories (Figure 5D). This meant that determining the contributions of various features to the two main principal components was significant, and the significance lay in the ability to identify features with the greatest differences between groups. On the basis of these findings, Variable Importance in Projection (VIP) score, which measured the contributes of each input features were calculated. And with the positive and negative values of feature loadings calculated by the PLS-DA model, we have assigned directionality to the VIP values, in which positive values with larger absolute value meant larger contributions to after intervention group and vice versa (Figure 5E). Then a ranked gene list was obtained and thus Gene Set Enrichment Analysis (GSEA) was conducted to evaluate pathway level changes from the perspective of machine learning. Last but not least, it was found that pathways like antigen process and presentation, as well as T cell receptor signaling pathway (Figure 5F) got significantly enriched (*P* value <0.05) by the gene list ranked by directional VIP score, while biological processes related to T-helper cells and biomolecules like IL-4 and IFN-γ were also enriched (Figure 5G).

### Directly and indirectly potential effects of TZD on regulating T cell in AR

With the combination of 16S rRNA gut microbiome metagenomics, serum transcriptomics and network target analysis, we have found that TZD potentially affected biological processes and pathways related to T cells in multiple ways. Among them, directly potential regulation through pathways represented by antigen process and presentation, together with indirect potential regulation through gut microbiome like Prevotella were focused in this research owing to their significant role (Figure 6A) on the basis of the combination of network target analysis and calculation of omics data. In details, on the one hand, TZD played a regulatory role in gut microbiome, influencing on certain microbiomes represented by Prevotella, which had been reported to have closely association with immune response^57,58^. Specifically, they were involved in a variety of T cell related biological processes, like T cell differentiation^59^, regulatory T cells process^60,61^ and other T cell regulation^62,63^. On the other hand, in MHCI pathway, TZD potentially affected TAP1/2, CALR, HSP70 and HSP90, as well as IFN γ which played an immune-proteasome role. Then through MHCI, TZD potentially affected T cell receptor signaling of CD8+ T cells and regulate the activation of NK cells. Besides, TZD had a potentially regulatory effect on CIITA/CREB/MHCII and HLA-DM, regulated CD4+ T cells and thus influenced downstream pathways and biological processes, like cytokine production and activation of other immune cells. Taking the relationship between gut microbiome and antigen process into consideration, there have already been some researches revealing their potential association. Research has shown that Prevotella primarily triggers Toll-like receptor 2, leading antigen-presenting cells to produce cytokines that favor Th17 polarization, such as interleukin-23 (IL-23) and IL-1^64^. Besides, enriched Faecalibacterium was observed in human leukocyte antigen (HLA)-B27+ individuals^65^ and a significant link between the Acinetobacter to Proteobacteria ratio and both the production of IL-13 and the likelihood of IL-13 production following allergen exposure was found^66^.

**Figure 6.**
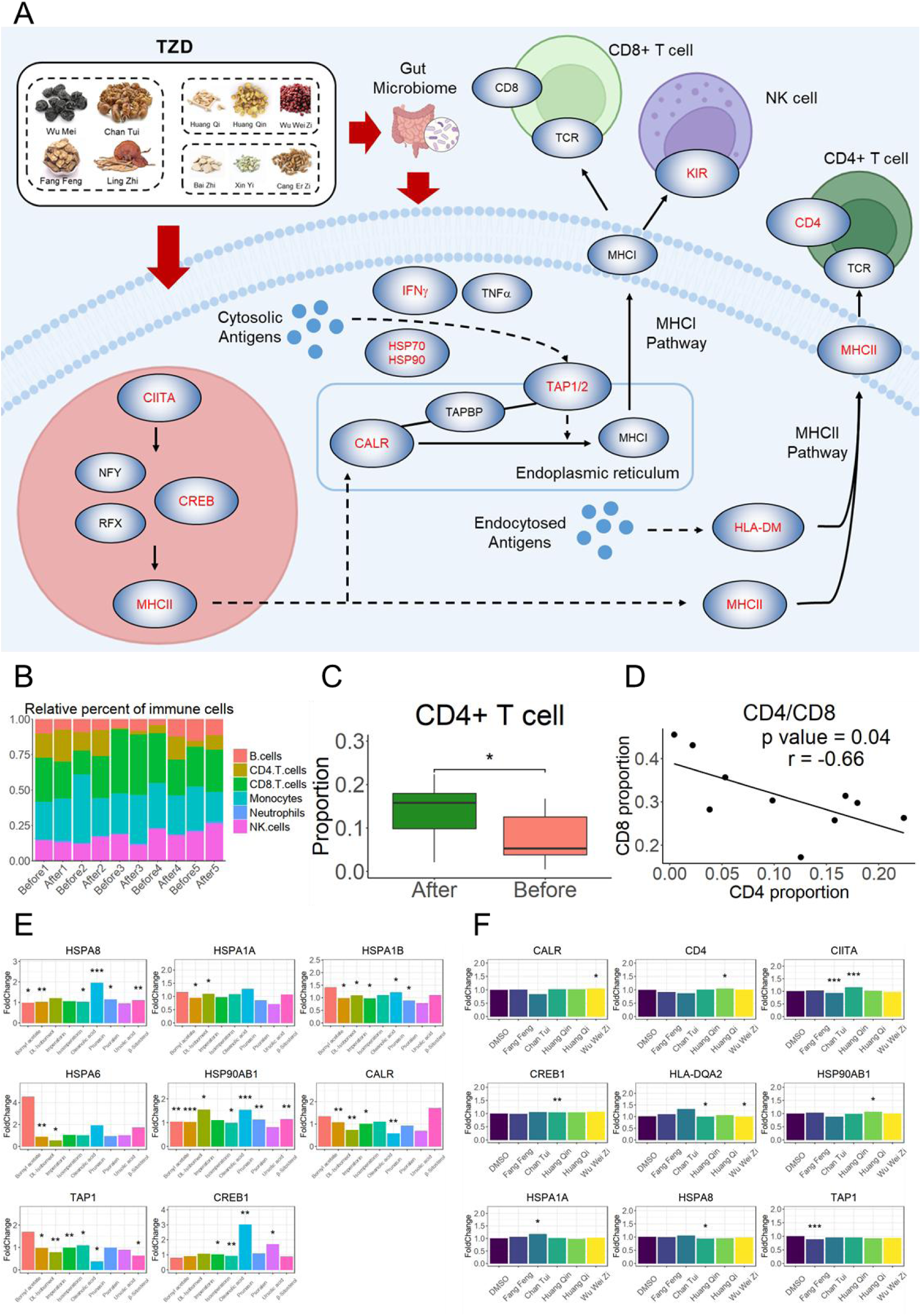
(A) Potential effects of TZD in antigen process and presentation on biomolecules and pathways, showing direct regulation on T cells through regulating antigen related process and indirect regulation on T cells through regulating gut microbiome. (B) Bar plot showing proportions of immune cells in different patients before and after the intervention of TZD. (C) Inferred proportion of CD4+ T cells before and after the intervention of TZD in AR patients. (D) Dot plot showing the increasing trend of CD4+ T cells and decreasing trend of CD8+ T cells in patients before and after the intervention of TZD. (E) Public validation based bar plot showing fold change and statistical significance of the regulatory effect of predicted key compounds on the expression of biomolecules related to the network target of TZD against AR. (F) Public validation based bar plot showing fold change and statistical significance of the regulatory effect of recorder herbs on the expression of biomolecules related to the network target of TZD against AR.

Further focusing on the downstream immune processes affected by TZD intervention, we utilized a deconvolution algorithm, CIBERSORT^67^, to infer changes in the proportions of immune cells from the serum transcriptomes before and after the intervention of TZD (Figure 6B). It was found that the proportion of CD4+ T cells showed a significantly increase after the intervention (Figure 6C), which to some extent confirmed our findings from the perspective of downstream outcomes. Besides, the proportion of CD8+ T cells and the proportion of CD4+ T cells had a negative relationship in AR patients before and after the intervention of TZD (Figure 6D). In addition, validation of regulation of TZD on antigen process related pathways and biomolecules were conducted based on several public datasets. Firstly, based on ITCM database^68^, which recorded expression of MCF-7 cell lines after the intervention of compounds from usual herbs, transcriptomics data of certain compounds of herbs in TZD were collected. After estimating the potential effects on antigen process and T-helper cell related biological process, 21 compounds belonging to herbs of TZD were kept for further statistical analysis. And it was found that 9 of them, like Ursolic acid and Oleanolic acid, had significant regulation on the predicted network target of TZD against AR (Figure 6E) under certain cell line conditions. Besides, on the basis of previous study^69^, the expression omics data of THP-1 cell line after the intervention of certain Jun herbs including Fangfeng and Chantui, as well as Chen herbs like Huangqin, Huangqi and Wuweizi were also collected. And on the predicted network target, they had different regulatory effects (Figure 6F). These public data based validations to some extent confirmed the potential intervention effects of TZD on these predicted targets. It also indicated the presence of synergistic effects involving multiple components and targets, and provided certain evidence for the selection of key compounds of TZD in the treatment of AR as well as the elucidation of JCZS.

## Discussion

As an allergic inflammation of the nasal mucosa characterized by symptoms such as nasal blockage, running nose, repeated sneezing, AR could seriously affect the quality of life of patients. TCM intervention has become an important way to effectively prevent and treat AR^2^. TZD, as a multi-component TCM formula, is an effective method for clinical intervention of AR, and its molecular mechanism needs to be further studied. Through integrating computational prediction, clinical multi-omics and experimental validation, this research uncovered the regulatory role of TZD against AR as well as identifying synergistic effects of herbs and compounds within TZD.

In this study, based on network target analysis, we constructed a multi-layer modular network. In this network, where immune-related modules played a dominant role, the potential intervention effect of TZD on AR was characterized. Guided by computational predictions, a single-arm clinical trial was initiated to validate the efficacy and safety of TZD in intervening AR. And the trial confirmed TZD’s effectiveness in alleviating symptoms, reducing IgE levels, and its preventive role against AR recurrence. Alongside the clinical trial, a comprehensive multi-omics clinical study was conducted, encompassing gut microbiome metagenomics and serum transcriptomics. In gut microbiome analysis, it was found that microbiomes like Firmicutes, Prevotella and Faecalibacterium decreased after the intervention of TZD, while the relative abundance increased in Firmicutes, Megamonas and Bacteroides. And in serum transcriptomics, pathways and biological processes related to immune response represented by antigen process and immune cell processes were significantly enriched. Based on the combination of computational analysis and experimental validation, the regulatory effect on immune response of TZD was found on direct effect on antigen process which regulate downstream immune process, as well as indirect regulation through gut microbiomes which has been reported to play regulate role on immune response in allergic diseases and other diseases^70–74^. Finally, regulatory effects on predicted important biomolecules in antigen process of vital compounds of TZD and herbs of TZD were verified on the basis of public omics data, which validated the potential influence of TZD on antigen process.

There are some limitations in our study. First, compounds of TZD were collected from the compositions of herbs recorded in database, which might have certain differences. Fortunately, algorithms applied in network target analysis could make up for the bias to some extent and still characterized the potential effects of TZD with representative compounds in a high precision. Second, our experimental validation has room for further improvement in both depth and breadth. This also represents a point worthy of focus in future research.

In conclusion, this work focused on deciphering the complex regulatory mechanisms of TCM against diseases. With the combination of network analysis, clinic trials and multi-omics technologies, we revealed the potential mechanism of TZD in the treatment of AR.

## Data Availability

All data produced in the present study are available upon reasonable request to the authors.
All data produced in the present work are contained in the manuscript.

## Abbreviations

TESS: Total Eye Symptom Score;
TNSS: Total Nasal Symptom Score;
TZD: Tuomin-Zhiti-Decoction;
AR: Allergic Rhinitis;
SAR: Seasonal Allergic Rhinitis;
RQLQ: Rhino conjunctivitis Quality of Life Questionnaire;
TCM: Traditional Chinese Medicine.

## Acknowledgments

This study was supported by the Anhui Province Traditional Chinese Medicine Science and Technology Research Project (202303a07020001) and Special project of the interpretation plan of the principles of traditional Chinese medicine of the State Administration of Traditional Chinese Medicine.

## Author Contributions

W.Z., B.W., Q.W., and S.L. designed the experiments. W.Z. and J.W. recruited patients and provided samples. B.W. and W.Z. performed multi-omics data analysis. The manuscript was drafted by B.W., S.L., W.Z. and critically revised by all the authors and discussed the results and commented on the manuscript by all the authors.

